# Hemispheric Asymmetry Defines Brain Aging: Five Reproducible NMF Modes Linked to Sex, Lifestyle, Transdiagnostic Genetic Risk, and Molecular Pathways

**DOI:** 10.64898/2025.12.26.25343034

**Authors:** Xiaobo Liu, Lang Liu, Zhou Le, Sanwang Wang, Zhao Zhang, Yong Han

**Affiliations:** Department of Psychiatry, Henan Mental Hospital, the Second Affiliated Hospital of Xinxiang Medical University, Xinxiang, China; McConnell Brain Imaging Centre, Montreal Neurological Institute, McGill University, Montreal, Canada; Department of Human Genetics, McGill University, Montreal, Canada; Peking University Sixth Hospital, Peking University Institute of Mental Health, NHC Key Laboratory of Mental Health (Peking University), Beijing, China; Department of Psychiatry, Renmin Hospital of Wuhan University, Wuhan, Hubei Province, China; Henan Key Lab of Biological Psychiatry, International Joint Research Laboratory for Psychiatry and Neuroscience of Henan, Xinxiang Medical University, Xinxiang, China; Henan Collaborative Innovation Center of Prevention and Treatment of Mental Disorder, Xinxiang, China; Institute of Brain Science, Henan Academy of Medical Sciences, Zhengzhou, China

**Keywords:** hemispheric asymmetry, cortical thickness, sex differences, lifestyle, transcriptomics, polygenic risk scores

## Abstract

Population aging heightens the burden of cognitive decline and brain disorders, yet trajectories of brain aging vary widely across individuals. Because the human brain is intrinsically lateralized, age-related shifts in hemispheric asymmetry may reveal latent aging subtypes that are masked by bilateral averages. Here, we derived reproducible and interpretable asymmetry-based brain-aging modes and validated their behavioral, genetic, and molecular signatures.

Using UK Biobank MRI, we computed cortical-thickness asymmetry across 68 Desikan–Killiany regions, transformed signed asymmetry into non-negative channels, and assembled a region-by-participant matrix. We then applied non-negative matrix factorization (NMF) to estimate spatial mode maps and participant-specific loadings, selecting the factorization rank by reconstruction-error elbow criterion (k = 13). Age associations were assessed with covariate-adjusted partial correlations controlling sex and handedness and corrected for multiple testing using false discovery rate (FDR). Generalizability was evaluated by projecting an independent cohort (Cam-CAN; n = 608) onto UK Biobank–derived spatial maps. We additionally tested sex differences, lifestyle/behavioral correlates, transdiagnostic polygenic risk score (PRS) coupling across 12 neuropsychiatric/neurodegenerative disorders, and imaging–transcriptomic pathway enrichment using Allen Human Brain Atlas expression and Metascape.

We identified five age-linked asymmetry modes that replicated directionally in Cam-CAN. Modes differed systematically by sex and displayed distinct lifestyle signatures spanning sleep, physical activity, alcohol intake, diet, device use, and smoking. Genetic coupling was mode-specific, with different modes aligning with distinct constellations of transdiagnostic PRS. Imaging–transcriptomic analyses further indicated mechanistic dissociability, implicating mitochondrial bioenergetics, antigen presentation, innate immune/inflammatory pathways, and synaptic/neurodevelopmental programs.

Hemispheric asymmetry decomposes into reproducible, mechanistically diverse aging modes that connect to modifiable behaviors and transdiagnostic genetic liability. This asymmetry-informed, mode-based framework advances subtype-oriented phenotyping of brain aging and provides a foundation for individualized risk stratification and mechanistic hypothesis generation.

## Introduction

Rising life expectancy has produced unprecedented population aging, elevating age-related cognitive decline and brain disorders to a pressing public-health concern^1^. Neurodegenerative change is a leading driver of disability and loss of independence, yet aging is biologically heterogeneous: some individuals retain near-youthful brain structure and cognition, whereas others exhibit accelerated decline^2^. The human brain is intrinsically lateralized in structure and function, and aging modulates this asymmetry^3^. Older adults often show reduced functional lateralization (e.g., HAROLD), and structural asymmetries display subtle, region-specific age effects^4^. Although mean effects are modest at the population level, between-person variability is large and prognostically meaningful^5^. Incorporating hemispheric asymmetry can therefore reveal “hidden” aging subtypes that bilateral aggregates may obscure^6^.

Brain aging is not a single axis but a multicomponent process unfolding across interacting networks, with distinct regional and tissue trajectories^7^. Collapsing aging into a single “brain-age gap” can mask biologically meaningful variation^8^. Data-driven decompositions—especially nonnegative matrix factorization (NMF)—yield sparse, additive, and interpretable spatial patterns with subject-level loadings while avoiding cancellation from mixed signs^9^. Prior work shows these components align with specific systems (e.g., frontoparietal cortical thinning, white-matter alterations) and display distinct genetic, lifestyle, and clinical correlates^10^. This pattern-based framework is particularly suited to capturing the complex effects of aging on structural asymmetry. In short, network-aware decomposition offers a principled lens to parse heterogeneous aging into separable, mechanistically informative modes^11^.

An asymmetry-informed approach tests whether left- versus right-dominant aging profiles entail distinct cognitive and clinical consequences. Elevated brain-age relates to poorer contemporaneous cognition, faster subsequent decline, greater risk of Alzheimer’s conversion, and higher mortality^12^, while lateralization anomalies are reported across multiple psychiatric disorders^13^. Linking asymmetry patterns to polygenic risk scores (PRS) and real-world behavior (e.g., hour-by-hour actigraphy) can differentiate susceptibility from resilience profiles^14^. Such integration clarifies intersections between normative aging variability and psychopathology, where transdiagnostic neurodegenerative pathways may operate^15^. It also supports risk stratification and highlights potentially modifiable lifestyle factors^16^. Overall, asymmetry provides a complementary dimension beyond the mere “rate” of aging^17^.

We compute MRI-derived asymmetry index (AI) across diverse structural measures, split signed AI into nonnegative channels, and assemble a region-by-subject matrix. Using NMF, we extract sparse, interpretable asymmetry factors and subject loadings, and replicate spatial patterns and projections in an independent cohort to establish stability. We then relate subtype membership and factor loadings to cognition, behavior, and the prevalence of 12 psychiatric disorders. Next, we test associations between aging-linked asymmetry patterns and multi-disease PRS, and evaluate links to daily activity phenotypes derived from 7-day wrist accelerometry. This lateralization-centric, pattern-based framework identifies asymmetry-defined aging subtypes, compares their behavioral/cognitive profiles, and assesses transdiagnostic clinical associations. Collectively, it advances individualized modeling of brain aging and bridges normative aging neuroscience with psychiatric epidemiology.

## Results

### Data-driven identification of aging modes and multidomain validation

Using NMF on the UK Biobank (UKB) cortical-thickness asymmetry matrix (regions × subjects; Desikan–Killiany 68 atlas), we decomposed the data into a regional weight matrix (W) and a subject loading matrix (H). Modes were retained on the basis of their partial correlation with age (two-sided tests, controlling sex and handedness; FDR across modes). This screening yielded five age-associated modes, which were subsequently interrogated across four external domains—cognition, lifestyle/behavior, polygenic risk for psychiatric/neurological disorders, and molecular pathways—each with domain-specific multiple-comparison control. Together, **Fig. 1** summarizes the analytic pipeline from NMF factorization to multidomain validation.

**Fig. 1.**
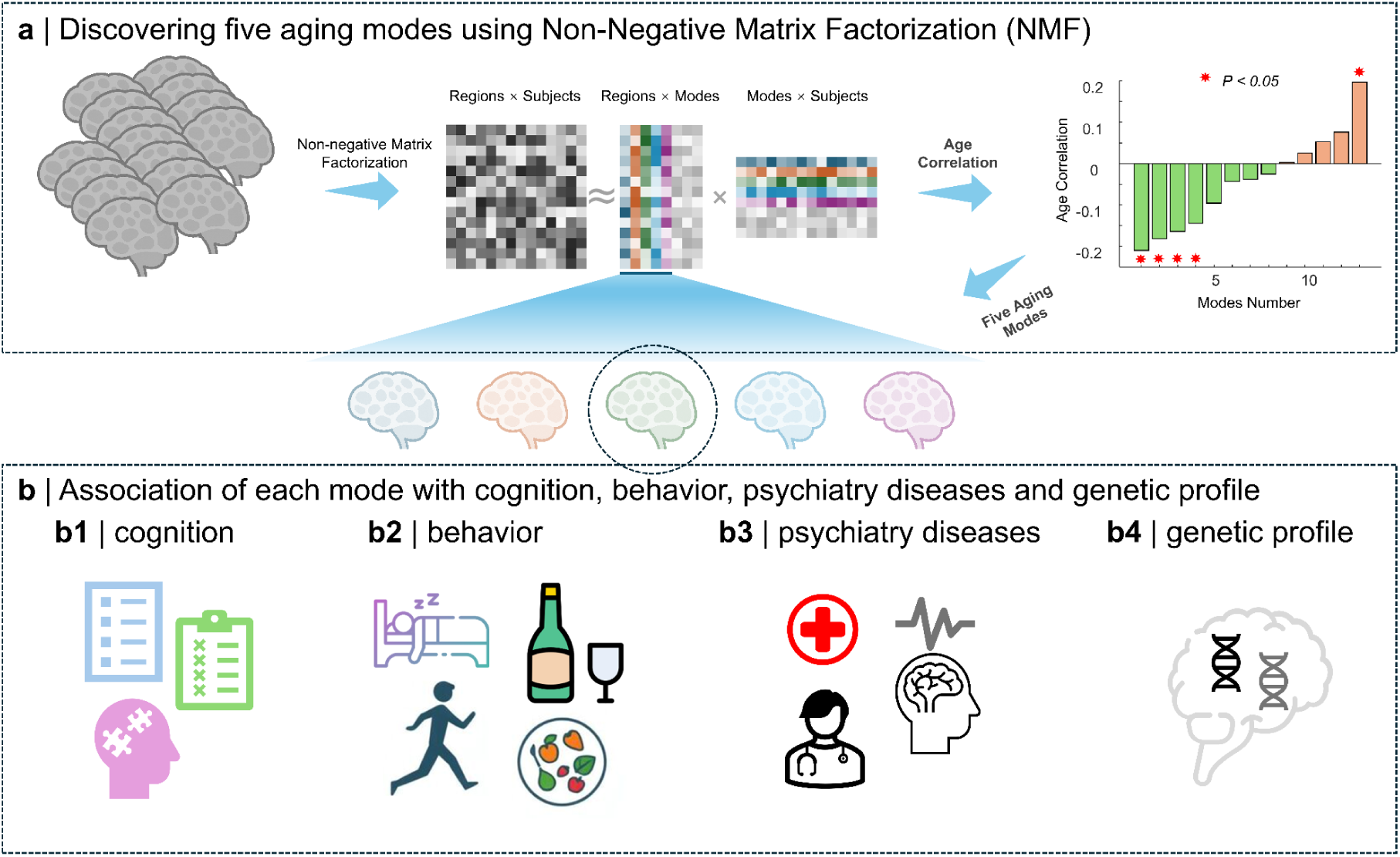
Schematic diagram of the study design. **(a)** Age-related latent brain patterns were identified using non-negative matrix factorization (NMF). The group “regions × subjects” matrix was decomposed into “regions × modes” and “modes × subjects,” and the optimal number of modes was determined by each mode’s correlation with age, yielding five distinct “brain-aging modes.” **(b)** Multidomain associations of each aging mode: **(b1)** cognition (word-cloud illustration highlighting language, sentence comprehension, semantic and autobiographical memory, working memory, etc.); **(b2)** behaviors (lifestyle measures including sleep, alcohol/substance use, physical activity, and diet); **(b3)** psychiatric disorders (radar-plot examples of association strength with conditions such as anxiety, insomnia, and depression); **(b4)** genetic profile (bubble/dot plots summarizing gene-level associations).

### Component-number optimization, age associations, and replication

We selected the NMF rank by evaluating reconstruction error across candidate mode numbers and applying an elbow criterion, which identified an optimal solution at *k* = 13 (Fig. 2a). Within this solution, five modes showed significant associations with age in UKB after FDR correction (Fig. 2b; FDR *q* < 0.05). To test generalizability, we fixed the UKB spatial maps and projected the independent Cam-CAN cohort (N = 608; 307 men/301 women; 54.19 ± 18.19 y) onto these bases to obtain out-of-sample loadings; the age–loading relationships replicated with matched directions and remained significant after FDR correction (Fig. 2c). The five exemplar cortical maps (Fig. 2d) demonstrated distinct spatial organizations. Quantitatively, age was negatively associated with Modes 1–4 and positively associated with Mode 5 (partial correlations controlling sex and handedness): Mode 1, *r* = −0.210; Mode 2, *r* = −0.181; Mode 3, *r* = −0.164; Mode 4, *r* = −0.145; Mode 5, *r* = 0.197 (all *p* < 0.05; Fig. 2e). These results indicate that asymmetry-linked brain aging comprises multiple latent trajectories with divergent age dependencies rather than a single monotonic process.

**Fig. 2.**
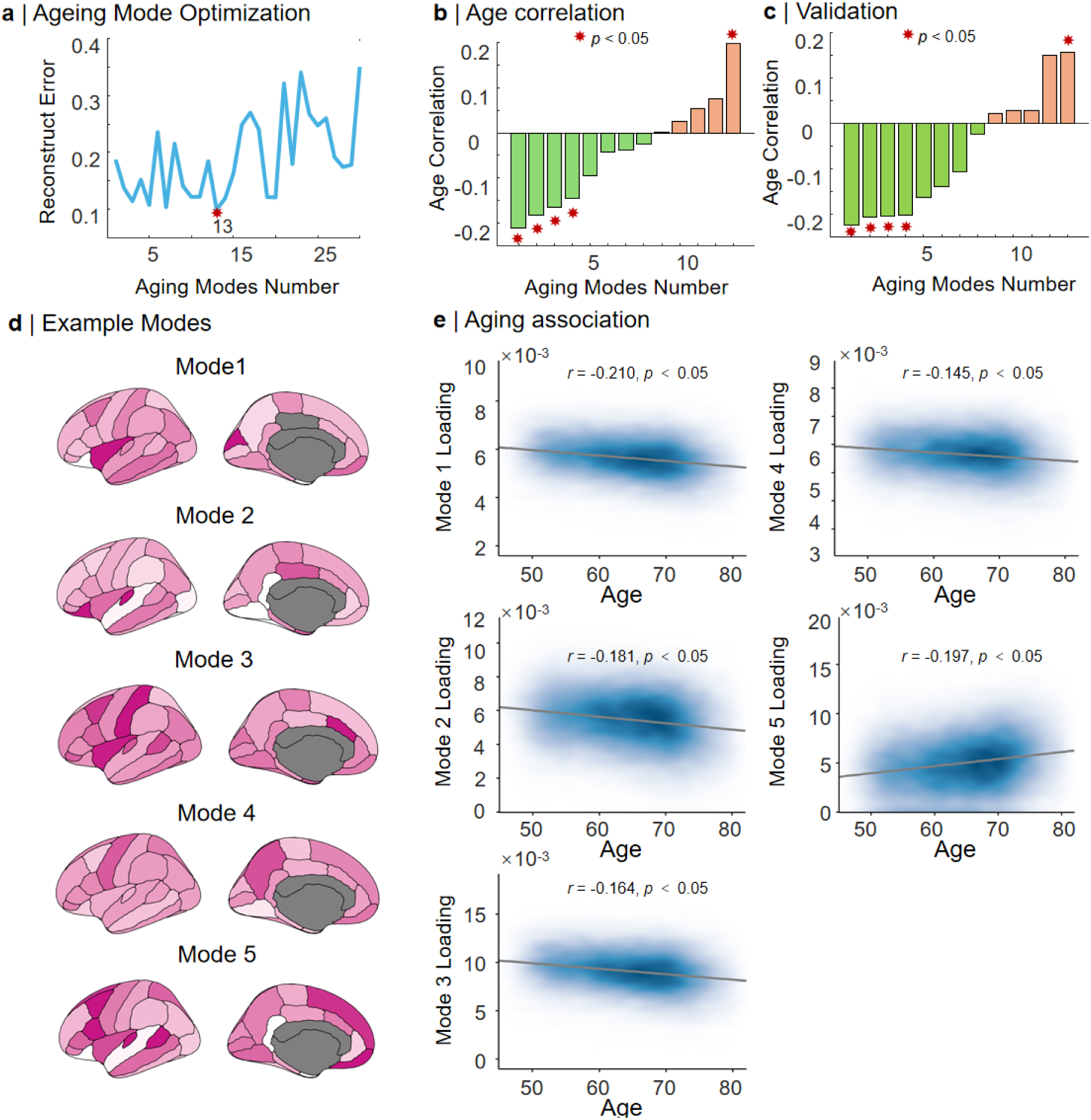
Determination of aging modes and validation of their associations with age. **a.** The number of aging modes is optimized by minimizing reconstruction error; the red asterisk marks the optimal number (example: 13). **b.** Correlations between each mode and age (bar plot; orange, positive; green, negative). Red asterisks denote statistical significance (p < 0.05). **c.** Validation in an independent dataset, with symbols and colors matching panel b. **d.** Representative cortical maps for Mode 1–Mode 5; color intensity reflects the weight/loading of each mode in the corresponding cortical region (darker indicates higher loading). **e.** Scatter–density plots with linear fits showing the association between age and the loadings of five example modes (Pearson correlation coefficient *r* and *p* value reported; shading indicates point density).

### Sex differences in expression of the five aging modes

Building on the replicated age effects, we tested whether mode expression differed by sex using two-sample *t*-tests (two-sided), adjusting for age and handedness and applying FDR correction across modes. All five modes exhibited significant sex differences (Fig. 3): Mode 1, *t* = −14.88; Mode 2, *t* = 16.25; Mode 3, *t* = −30.64; Mode 4, *t* = 20.66; Mode 5, *t* = 25.41 (all FDR-corrected *p* < 0.05). The kernel-density and jitter/rug visualizations further show distributional shifts, indicating that sex is a major source of inter-individual heterogeneity in asymmetry-based aging profiles and should be modeled explicitly in downstream analyses.

**Fig. 3.**
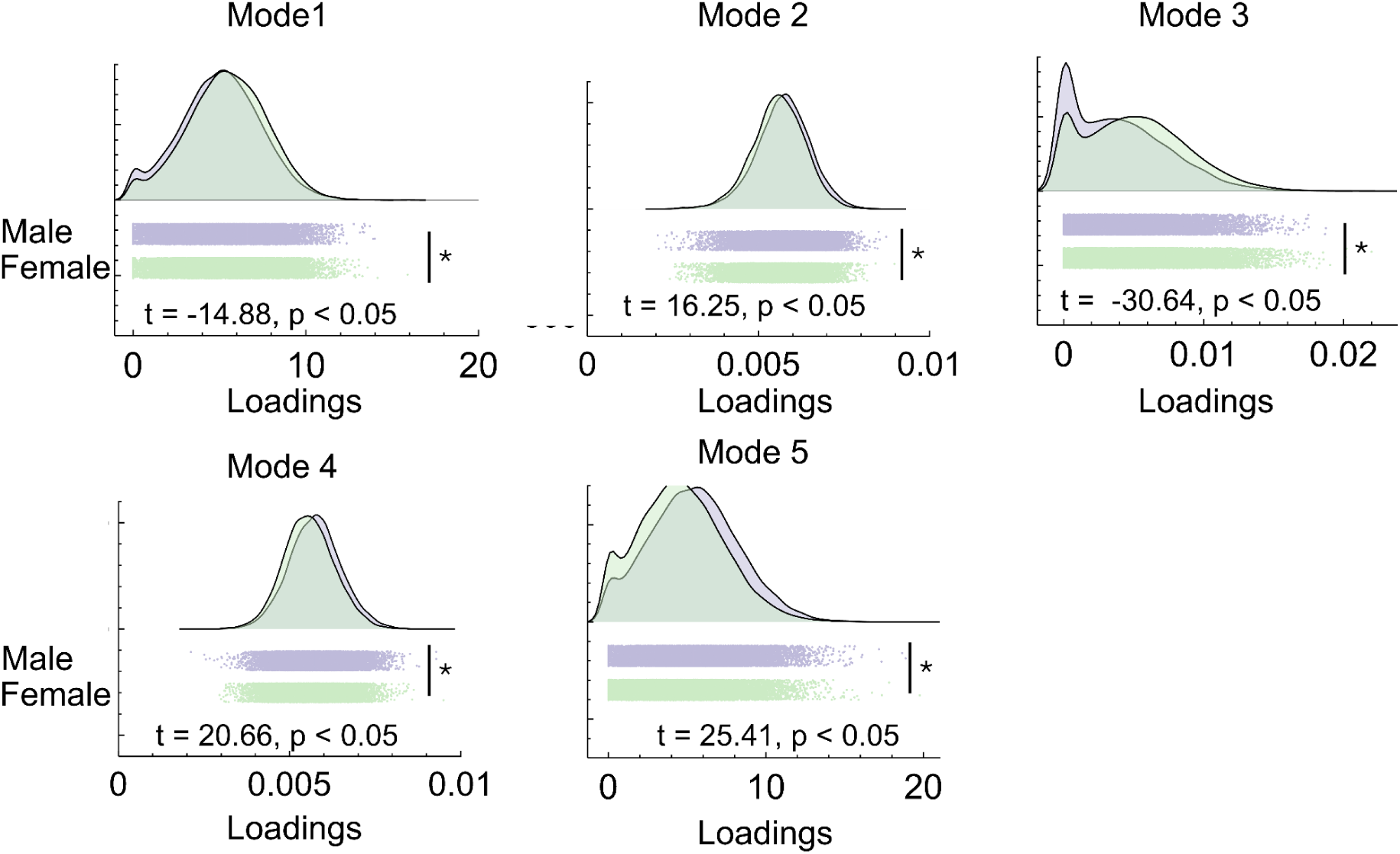
Sex differences across the five aging modes (Mode 1–Mode 5). Each panel compares male and female distributions of individual loadings: kernel density curves above (black outlines) and sex-colored rug/jitter plots below; the x-axis denotes mode loadings. Panel insets report two-sample *t* statistics and *p* values; asterisks at right indicate statistical significance (FDR *q* < 0.05). All five modes show significant sex differences, with the direction evident from the distributions and *t* values.

### Lifestyle and behavioral phenotypes exhibit mode-specific association signatures

To characterize real-world correlates of the five aging-related asymmetry modes, we tested associations between individual mode loadings and a broad panel of lifestyle/behavioral phenotypes using Pearson correlations. Multiple testing was controlled using false discovery rate (FDR) correction, and results were summarized as −log10(*p*) (Fig. 4; FDR *q* < 0.05). Across the full phenotype set, each mode displayed a distinct pattern of significant associations—an interpretable “fingerprint”—rather than a uniform set of correlates shared across modes. Notably, significant associations spanned diverse behavioral domains, including sleep/circadian rhythm, physical activity, diet, alcohol consumption, electronic device use, and smoking-related measures, underscoring pronounced heterogeneity in behavioral sensitivity across the asymmetry-defined aging trajectories.

**Fig. 4.**
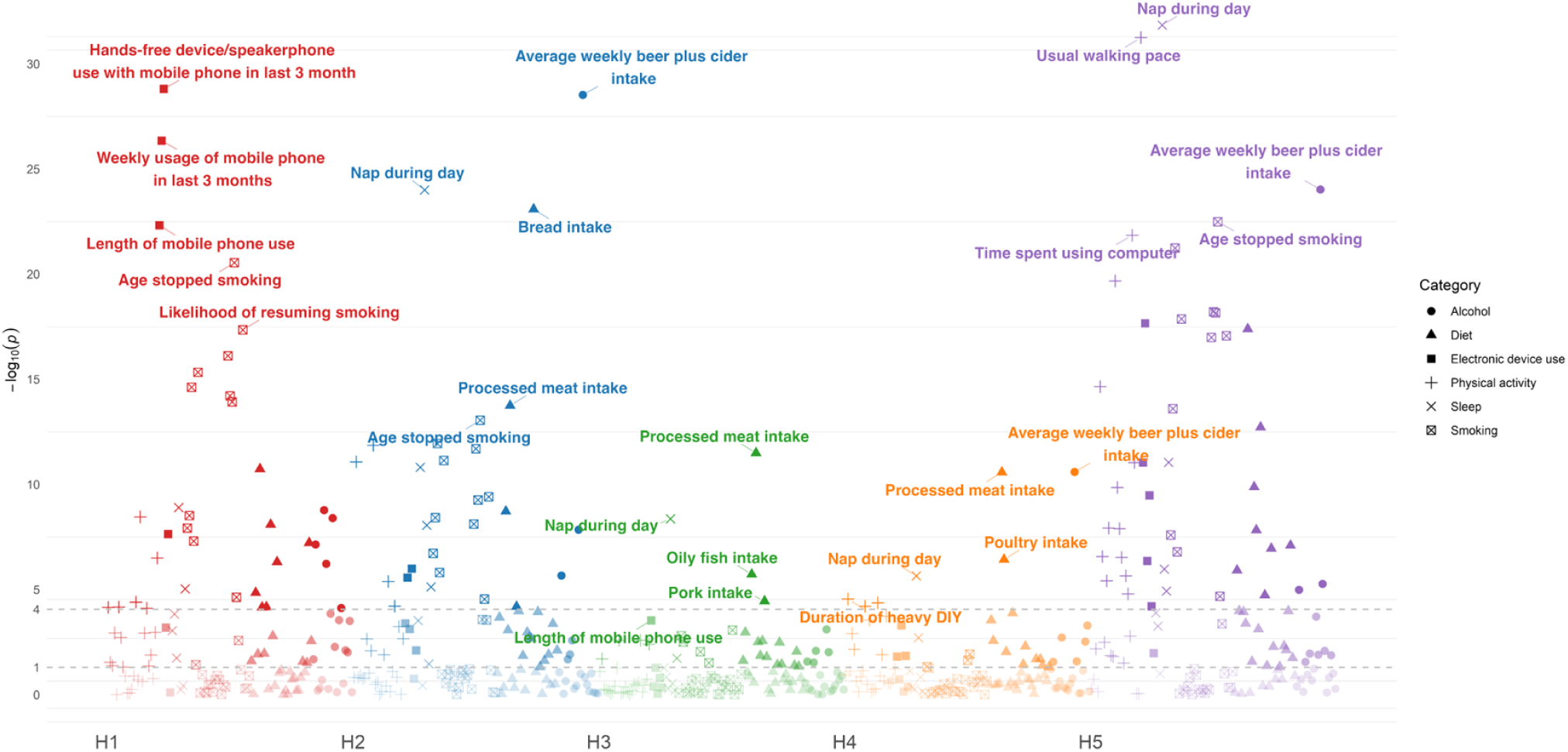
The statistical significance of associations between a range of lifestyle factors and five modes. The x-axis indicates the modes, while the y-axis shows significance as (−log_10_(*p*)); gray dashed lines mark selected significance thresholds.

Among the most prominent signals highlighted in Fig. 4, several phenotypes repeatedly emerged as FDR-significant, albeit with mode-dependent strength and/or direction. Sleep-and rhythm-related measures were prominently represented, with daytime napping (“nap during day”) showing robust associations in multiple modes, suggesting that variation in sleep propensity or circadian regulation co-varies with specific asymmetry aging patterns. Physical-activity proxies also featured strongly: usual walking pace (“usual walking pace”) reached high significance for select modes, consistent with the view that cardiorespiratory fitness and habitual mobility are tightly coupled to brain aging phenotypes but may map preferentially onto particular lateralized network patterns. Alcohol-related behaviors contributed additional mode-specific signals, with average weekly beer/cider intake (“average weekly beer plus cider intake”) repeatedly significant across modes, indicating that alcohol exposure is not merely related to global aging but may align with specific asymmetry trajectories. Diet-related phenotypes were also evident; processed-meat intake (“processed meat intake”) recurred among significant associations, supporting a link between dietary patterns and asymmetry-linked structural variation that may reflect broader cardiometabolic or inflammatory pathways. Contemporary digital behavior showed mode-selective associations: mobile-phone use duration/frequency (e.g., “length of mobile phone use,” “weekly usage of mobile phone in last 3 months,” and hands-free/speakerphone use) was particularly salient for certain modes, suggesting that device-use metrics track behavioral/lifestyle constellations that intersect with asymmetry aging patterns, though interpretation remains vulnerable to confounding and reverse causality. Finally, smoking-related traits (e.g., age stopped smoking and likelihood of resuming smoking) were significant for specific modes, indicating that smoking history and relapse propensity differentially relate to distinct asymmetry aging dimensions.

Collectively, these results indicate that asymmetry-derived aging modes capture more than a pure chronological aging effect. Instead, they encode separable brain-aging trajectories that are differentially coupled to lifestyle and behavioral exposures, implying that distinct lateralized aging patterns may reflect different environmental sensitivities, compensatory behavioral profiles, or correlated health states.

### Transdiagnostic PRS associations reveal distinct genetic susceptibility profiles across modes

We evaluated whether asymmetry aging modes are differentially coupled to inherited liability for major neuropsychiatric and neurodegenerative disorders by testing associations between mode loadings and polygenic risk scores (PRS) for 12 conditions (PD, insomnia, SCZ, MDD, anxiety, BP, AUD, PAU, PTSD, ALS, LBD, AD). Associations were estimated using partial correlation/regression models controlling for age, sex, and handedness, and significance was determined after FDR correction across all mode × PRS tests (Fig. 5; FDR *q* < 0.05). Radar plots revealed that each mode expressed a distinct PRS association pattern, supporting genetic separability among the lateralized aging dimensions.

**Figure 5.**
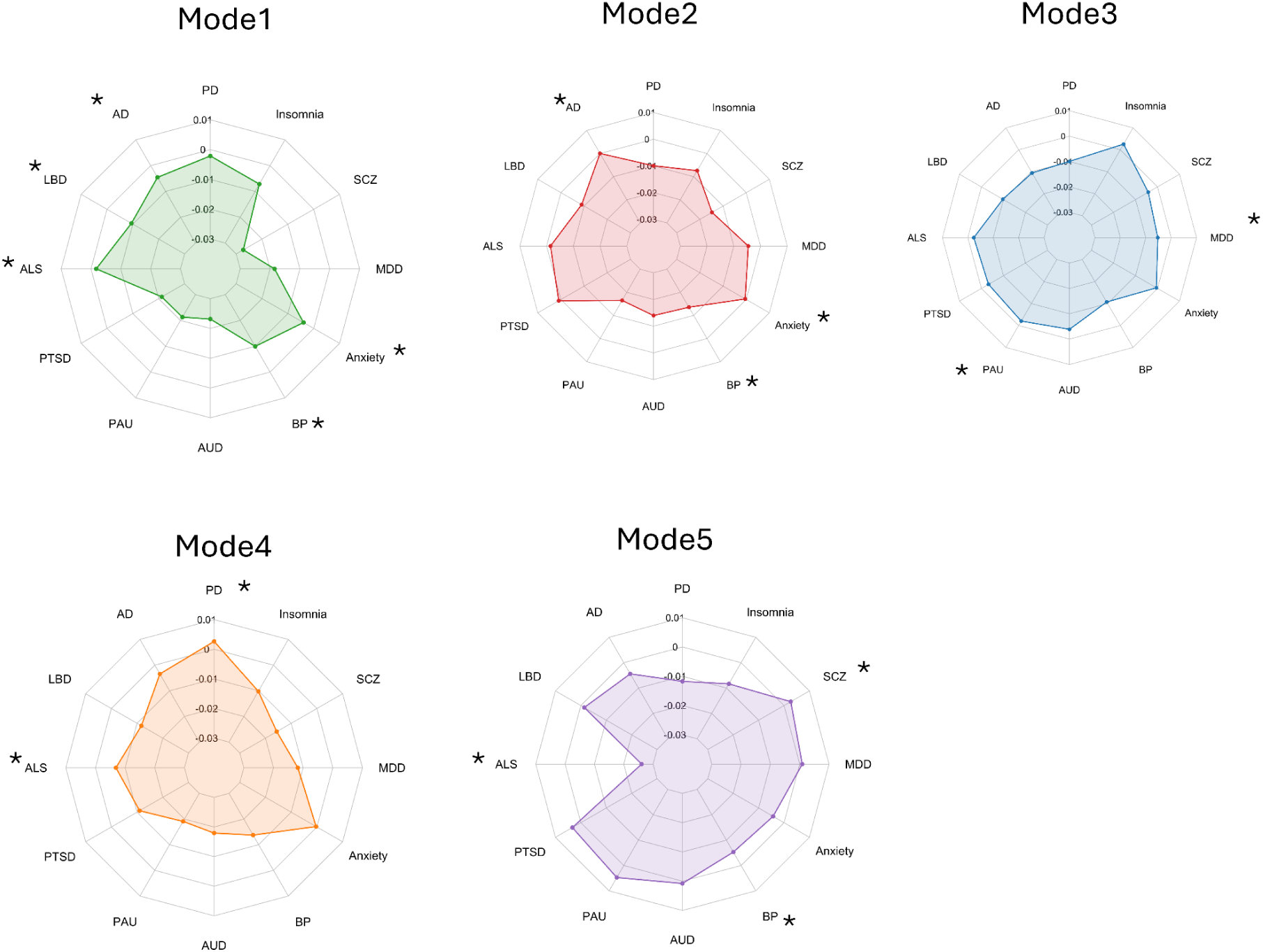
Radar charts showing the associations between five latent modes and polygenic risk scores (PRS) for 12 neurological/psychiatric disorders. Colors distinguish modes. Concentric circles indicate the effect-size scale. Asterisks mark statistically significant associations after multiple-comparison correction. PD: Parkinson’s disease; SCZ: schizophrenia; MDD: major depressive disorder; BP: bipolar disorder; AUD: alcohol use disorder; PAU: problematic alcohol use; PTSD: post-traumatic stress disorder; ALS: amyotrophic lateral sclerosis; LBD: Lewy body dementia; AD: Alzheimer’s disease. * means significance after FDR.

Summarizing the FDR-significant associations indicated by asterisks in Fig. 5, Mode 1 showed significant coupling with PRS for Alzheimer’s disease (AD), Lewy body dementia (LBD), amyotrophic lateral sclerosis (ALS), anxiety, and bipolar disorder (BP) (all FDR *q* < 0.05), suggesting that this asymmetry trajectory aligns with shared liability spanning neurodegeneration and affective/anxiety-related genetic risk. Mode 2 was significantly associated with AD, anxiety, and BP PRS (FDR *q* < 0.05), indicating a partially overlapping but narrower genetic profile relative to Mode 1. Mode 3 displayed significant associations with major depressive disorder (MDD) and problematic alcohol use (PAU) PRS (FDR *q* < 0.05), implying that this mode may reflect a lateralized aging dimension preferentially linked to mood and addiction-related liabilities. Mode 4 was significantly associated with Parkinson’s disease (PD) and ALS PRS (FDR *q* < 0.05), highlighting a genetic signature enriched for motor/neurodegenerative risk. Mode 5 exhibited significant associations with schizophrenia (SCZ), BP, and ALS PRS (FDR *q* < 0.05), suggesting coupling to a distinct transdiagnostic liability profile spanning psychosis-related risk, bipolar liability, and a neurodegenerative component.

Taken together, these findings demonstrate that asymmetry-defined aging modes are not only phenotypically heterogeneous but also genetically differentiated: different lateralized aging trajectories appear to share distinct components of common-variant liability with specific neuropsychiatric and neurodegenerative disorders. This transdiagnostic coupling supports the interpretation that the modes index separable biological pathways rather than being purely statistical decompositions of a single underlying aging process.

### Imaging–transcriptomic anchoring reveals mechanistic heterogeneity in pathway enrichment across modes

To obtain molecular interpretations for the spatially distributed asymmetry aging modes, we integrated neuroimaging patterns with cortical transcriptomics. Specifically, we mapped gene-expression profiles from the Allen Human Brain Atlas (AHBA) onto the Desikan–Killiany parcellation and correlated each mode’s regional weight map with regional gene-expression levels. Genes whose spatial expression profiles significantly aligned with a given mode were then submitted to Metascape for pathway enrichment analysis, using an FDR threshold of 5% to define significant enrichment (Fig. 6; FDR *q* < 0.05). Results were visualized as bubble plots reporting GeneRatio, hit-gene counts, and −log10(*p*), alongside term networks summarizing functional clusters.

**Fig. 6.**
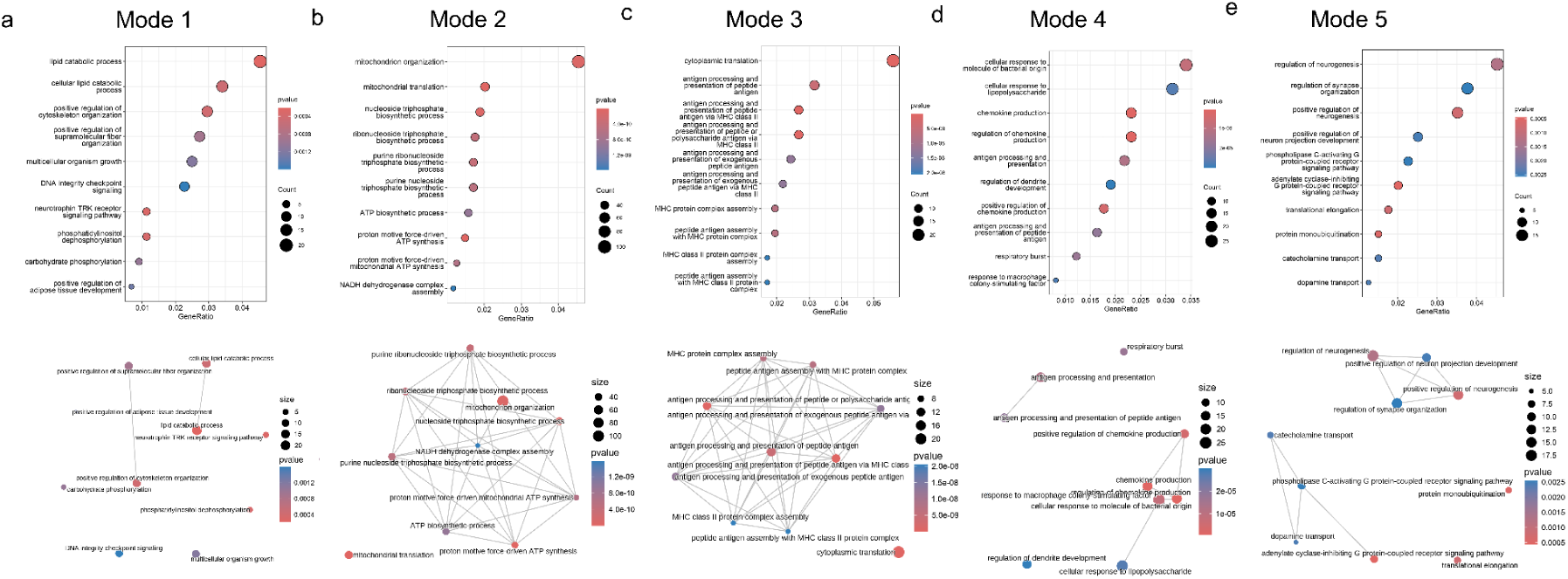
Functional enrichment results for five modes. The top panels show bubble plots, with the x-axis indicating the gene ratio; bubble size represents the number of hit genes, and color encodes significance (−log_10_(*p*)). For each mode, the top enriched terms are listed. The bottom panels display the corresponding term networks, where nodes denote significantly enriched biological processes/pathways, node size scales with gene count, and edges indicate shared genes between terms. Proximal nodes form functional clusters, providing an intuitive overview of the principal biological themes in each mode.

The enrichment profiles differed markedly across modes, implying distinct biological programs underlying different asymmetry-defined aging trajectories. Mode 2 showed strong enrichment for mitochondrial and energy-metabolism processes—such as electron transport chain activity, oxidative phosphorylation, ATP synthesis, and NADH dehydrogenase complex assembly—pointing to a mode that may be particularly related to bioenergetic efficiency and mitochondrial function. Mode 3 was enriched for antigen processing and presentation pathways, including MHC class I complex assembly and antigen presentation, implicating immune surveillance and inflammation-related molecular processes. Mode 4 was enriched for innate immune and inflammatory response programs, including responses to bacterial-derived molecules (e.g., LPS), regulation of chemokine production, and inflammatory activation cascades, consistent with a distinct immuno-inflammatory axis. Mode 5 exhibited enrichment for neurogenesis and synaptic signaling regulation, including terms related to neuronal development, modulation of synaptic transmission, and receptor-mediated signaling (including GPCR-related pathways), suggesting a mode potentially linked to synaptic plasticity and neural remodeling. Mode 1 showed enrichment for broad cellular signaling and regulatory processes (e.g., phosphorylation-dependent signaling cascades and regulation of cellular processes), consistent with a lateralized aging dimension reflecting coordinated modulation of signaling networks rather than a single narrowly defined pathway.

Overall, the imaging–transcriptomic analyses provide convergent evidence that the asymmetry-derived aging modes are mechanistically heterogeneous at the molecular level. Combined with the demographic, behavioral, and PRS findings, these results support the interpretation that the identified modes are not spurious statistical components but represent biologically meaningful and partially dissociable aging-related processes—spanning mitochondrial energetics, immune/inflammatory pathways, and synaptic/neurodevelopmental programs—each contributing in a distinct manner to individual variability in lateralized brain aging.

## Discussion

This study leveraged cortical thickness asymmetry as an entry point and used NMF to extract five interpretable aging modes from a large “regions × participants” matrix, yielding convergent external validity across cognition, lifestyle, cross-disorder polygenic risk, and molecular profiles. This network- and pattern-based approach aligns with prior evidence that brain aging is multicomponent, while establishing hemispheric asymmetry as an independent dimension for delineating aging subtypes. Owing to the nonnegativity and sparsity of NMF, lateralized signals are not canceled by mixed signs, enhancing interpretability and laying a methodological foundation for subsequent mode-based individualized assessments.

Hemispheric asymmetry is a fundamental organizational feature of the human brain that relates to specialization, efficiency, and vulnerability, and it can reveal lateralized aging signals that may be partially obscured when collapsing across hemispheres^18^. Non-negative matrix factorization provides a “parts-based” decomposition that often yields sparse and interpretable components, which is advantageous for mapping high-dimensional cortical patterns into mechanistically testable modes^19^. Our emphasis on individualized “mode loadings” also aligns with contemporary brain-aging and computational psychiatry work highlighting substantial inter-individual heterogeneity, motivating dimensional and normative approaches beyond average case–control contrast^20–22^.

We identified five modes that were significantly associated with age and replicated their directional associations in an independent sample. Most modes decreased with advancing age, whereas a minority increased, indicating that aging does not proceed “at the same speed in the same direction” but unfolds along distinct network trajectories. By fixing the UKB spatial maps and projecting individual loadings in an external dataset, we provide cross-cohort–transportable “asymmetry aging primitives.” The age–loading relationships and the spatial topology of these modes (e.g., fronto-parietal systems) are consistent with known vulnerable networks, strengthening biological plausibility.

The coexistence of decreasing and increasing asymmetry-loadings is consistent with cognitive-neuroscience models proposing that aging can involve both reduced hemispheric specialization and compensatory reconfiguration, depending on system demands and reserve^23^. Large-scale brain-aging studies similarly indicate that “apparent aging” is not a single axis but reflects multiple partially independent biological processes, supporting the rationale for decomposing aging into distinct modes^21,24,25^. Cross-cohort transportability is particularly valuable in population imaging where scanner/protocol differences and sampling can inflate spurious effects; UK Biobank-scale resources provide an essential foundation for robust discovery, while independent cohorts help validate generalization^26,27^.

All five modes exhibited significant sex differences, indicating a sex-specific expression profile of asymmetry-related aging that resonates with the long-term shaping of brain lateralization and aging trajectories by sex hormones, immune processes, and lifestyle. Unlike prior work focusing on single regions or global thickness, we demonstrate sex effects at the “mode level,” underscoring the need to incorporate sex a priori into risk stratification and intervention design, and to further examine the mediating roles of menopausal status, gene-by-sex interactions, and cardiometabolic indices in mode expression.

Sex differences in brain organization have been repeatedly observed across large neuroimaging datasets and meta-analytic evidence, spanning structural connectivity and regional morphometry, making it plausible that asymmetry-based aging phenotypes will also show sex-biased expression^28,29^. These results argue for sex-stratified (or at minimum sex-calibrated) reference models when interpreting individual deviations in asymmetry loadings, especially if the goal is risk stratification or precision prevention.

With respect to lifestyle, distinct modes showed robust associations with daytime napping, walking pace, alcohol type/frequency, mobile-phone use duration, and processed-meat intake. These results not only recapitulate established influences of physical activity, sleep, and drinking on brain aging but also map contemporary behavioral and dietary factors to specific asymmetry aging modes, pointing to “modifiable lifestyle–asymmetry network” pathways. Heterogeneous sensitivity of modes to environmental inputs likely reflects differences in network plasticity. Testing reversibility and dose–response relationships of mode loadings within a wearable, longitudinal framework will directly inform target selection for secondary prevention^25^.

Physical activity has experimental and longitudinal support for benefiting memory-relevant brain systems (e.g., hippocampal plasticity), providing a biologically grounded pathway through which “walking pace/fitness” proxies could relate to mode expression^30^. Sleep is mechanistically linked to brain homeostasis and metabolite clearance, offering plausible routes by which chronic sleep disruption or altered sleep architecture could shape structural aging phenotypes^31^. Alcohol exposure has been associated with adverse brain outcomes in large cohort analyses, consistent with the directionality observed for some lifestyle–mode associations^32^. Dietary-pattern evidence (e.g., MIND-style patterns) supports links between nutritional quality and cognitive/brain outcomes, suggesting that food-related signals may reflect broader cardiometabolic–neuroinflammatory pathways rather than single items alone^33^. By contrast, digital technology measures (e.g., mobile-phone duration) often show small, context-dependent associations and are vulnerable to confounding and reverse causality; interpreting these signals will require careful longitudinal and causal designs^34^. Accordingly, causal-inference strategies—including Mendelian randomization where assumptions hold—will be important for determining whether lifestyle variables are drivers, correlates, or consequences of altered asymmetry-defined aging trajectories^35^.

Cross-disorder PRS analyses revealed mode-specific positive and negative associations with twelve neuropsychiatric PRSs, with bipolar disorder (BP) PRS reaching multiple-comparison–corrected significance for several modes. This extends the literature on “psychiatric disorders with lateralization anomalies” from single-disorder/single-metric observations to a system-level mapping between cross-disorder genetic risk and asymmetry aging modes. Within a developmental–aging continuum, asymmetry modes linked to BP-PRS in adulthood may reflect nascent vulnerability already emergent in adolescence—manifested as subtle cortical thickness and structural covariance alterations—and closely tied to hemispheric specialization of executive and emotion-regulation networks. Thus, genetic liability for BP and related conditions may not only elevate disease risk but also bias hemisphere-specific aging trajectories in adulthood, offering critical clues for developmental precision psychiatry.

Polygenic risk score analysis is now a standard framework for summarizing common-variant liability, but interpretation depends on discovery-sample composition, ancestry portability, and trait architecture; these factors should be explicitly considered when translating PRS–brain links across cohorts^36,37^. The observed cross-disorder patterning is consistent with evidence for shared genetic risk across major psychiatric conditions, supporting transdiagnostic models in which overlapping liabilities express in distributed brain systems^38^. The specific PRSs highlighted here are grounded in large GWAS efforts for bipolar disorder, schizophrenia, insomnia, and Alzheimer’s disease, each implicating biologically coherent pathways that could plausibly intersect with asymmetric cortical aging^39–41^. Clinically, these findings motivate testing whether asymmetry-mode profiles improve prediction of symptom trajectories or treatment response beyond traditional risk measures, while ensuring rigorous calibration and avoiding ancestry-related inequities^22,25,42,43^.

Molecular enrichment further uncovered differentiated biological pathways underlying each mode, prominently including mitochondrial energy metabolism, protein processing and immune pathways, and synaptic/neurodevelopmental processes. Aligning “spatial asymmetry modes” with “molecular function modules” suggests that aging across hemispheres/networks does not share a single pathway. This multilayer correspondence—spanning molecules, networks, and behavior—provides a clear map for druggable and lifestyle-modifiable targets and offers a basis for monitoring pathway-specific responsiveness in intervention trials.

The prominence of mitochondrial, proteostasis, and immune-related terms is broadly consistent with canonical hallmarks of aging, supporting the plausibility that mode-specific asymmetry captures separable biological aging programs rather than a unitary process^44^. The convergence of immune–synaptic biology is also notable given strong evidence that complement-mediated synaptic remodeling is implicated in severe mental illness risk, offering one mechanistic bridge between PRS findings and molecular enrichment^45^. Imaging–transcriptomic anchoring using the Allen Human Brain Atlas provides a principled route to translate macroscale spatial patterns into molecular hypotheses, though interpretation must account for spatial sampling and donor limitations^46^. Prior work shows that correlated gene-expression patterns align with functional network organization, supporting the conceptual link between network-like spatial modes and transcriptomic gradients^47^. Because best practices for linking brain-wide gene expression and neuroimaging emphasize sensitivity analyses and methodological rigor, future work should replicate enrichment under alternative parcellations, mapping strategies, and control models^48^. Finally, pathway tools such as Metascape enable interpretable summaries of systems-level results, but triangulation with single-cell resources and orthogonal omics will strengthen causal specificity for intervention targeting^49^.

### Limitations

While this study provides valuable insights into the role of hemispheric asymmetry in brain aging, several limitations should be considered. First, the cross-sectional nature of the data restricts our ability to infer causal relationships between the identified aging modes and the observed behavioral, genetic, and molecular associations. Second, although the use of a large, well-phenotyped cohort (UK Biobank) enhances the generalizability of our findings, the study predominantly includes individuals of European ancestry. This may limit the applicability of the results to populations with different genetic backgrounds. Additionally, while we controlled for several demographic and lifestyle factors, other unmeasured variables, such as socioeconomic status, environmental exposures, and mental health history, could further influence brain aging. Future research is needed to explore the temporal dynamics, ethnic diversity, and broader environmental factors influencing hemispheric asymmetry and brain aging.

## Conclusion

In conclusion, leveraging cortical-thickness asymmetry in a large, well-phenotyped cohort, we identified five reproducible and biologically interpretable aging modes via NMF that generalize to an independent sample and show robust associations with age, sex, modifiable lifestyle factors, transdiagnostic polygenic risk, and mode-specific molecular pathways, thereby establishing hemispheric asymmetry as a substantive axis of neurobiological heterogeneity in aging. These results move beyond single-scalar “brain age” toward subtype-oriented representations that capture qualitative differences in how the brain ages (e.g., left- vs right-dominant trajectories) and offer candidate markers for risk stratification and precision prevention. While cross-sectional design and ancestry focus limit causal inference and generalizability, the framework is scalable and readily extensible to longitudinal, multimodal, and ancestrally diverse datasets, providing a foundation for testing whether behavioral or biological interventions can modulate asymmetry-defined aging trajectories.

## Method

### Data description

This research was conducted using the UK Biobank (UKB) resource under ethical approval from the North West Multi-Centre Research Ethics Committee as a Research Tissue Bank. The UK Biobank study protocol is publicly available (https://www.ukbiobank.ac.uk/), and all participants provided written informed consent for data collection and linkage. UK Biobank is a large-scale biomedical cohort comprising 502,467 individuals aged 37–72 years, recruited in the United Kingdom between 2006 and 2010. The resource includes extensive demographic, biological, lifestyle, environmental, physical measurement, mental health, and imaging data.

The demographics were shown in **S-Table 1**.

### MRI data quality control

T1-weighted MRI scans from the UKB were minimally preprocessed and subjected to cortical reconstruction in FreeSurfer (v6.0.1) using standardized pipelines^50^. The cerebral cortex was parcellated into 68 regions according to the Desikan–Killiany atlas, and inter-subject alignment was performed via surface-based registration of the T1-weighted images. Mean cortical thickness (CT), as a macrostructural MRI-derived phenotype, was then extracted and standardized for downstream analyses.

### Structural asymmetric pattern extraction

This study focuses on cortical thickness asymmetry, quantified by the asymmetry index (AI), defined as AI=L−R(L+R)/2, where L and R denote thickness in homologous left and right cortical regions, respectively. After computing AI for each region and participant, all asymmetry values were assembled into a region-by-subject data matrix. To extract interpretable asymmetry patterns, we split AI into two nonnegative channels (positive and negative components) and applied nonnegative matrix factorization (NMF) to decompose the data into a “pattern” matrix (regional weights) and an “individual loading” matrix (participant-wise expression of each pattern). To enhance interpretability, columns of the pattern matrix were L1-normalized (unit-sum weights), and sparsity constraints were imposed on loadings to encourage a small number of dominant patterns per individual. NMF was repeated across multiple candidate ranks (e.g., 2–20) with multiple random initializations, and the optimal rank was selected based on reconstruction error.

### Ageing effect on asymmetric pattern extraction

To assess aging effects, we quantified the association between individual NMF loadings and age (correlation) with false discovery rate (FDR) correction, including sex and handedness as covariates. Then we ranked components from negative to positive effect sizes. To evaluate robustness, we performed an out-of-sample reconstruction in an independent cohort: fixing the spatial patterns learned in UKB, we projected each participant in the Cambridge dataset ^51^ onto these bases to obtain subject-specific loadings and repeated the age–loading correlation analysis. We analyzed data from 608 healthy adults (307 men, 301 women; mean age = 54.19 ± 18.19 years) from the Cambridge Centre for Ageing and Neuroscience (Cam-CAN) ^51^, replicated the UKB MRI preprocessing pipeline, and extracted cortical thickness for each participant.

### Sex difference on asymmetric pattern extraction

We further conducted two-sample (independent) t-tests to assess sex differences in individual loadings across patterns, controlling for age and handedness as covariates, with false discovery rate (FDR) correction

### SNP Genotyping

Saliva-derived DNA samples were genotyped for approximately 550,000 single nucleotide polymorphisms (SNPs) ^52^ using the Illumina Human660W Quad BeadChip^53–55^. Genotyping data underwent preprocessing and quality control to ensure optimal imputation accuracy, leveraging the “imputePrepSanger” pipeline. The Human 660W-Quad_v1_A-b37-strand reference panel was employed for alignment and strand orientation to ensure consistency across datasets (available at: https://hub.docker.com/r/eauforest/imputeprepsanger/).

### Genotyping Quality Control and Imputation

Rigorous quality control (QC) procedures were implemented utilizing PLINK 1.9, adhering to established best practices for genome-wide association studies (GWAS). QC protocols included heterozygosity analysis via the PLINK –indep-pairwise command (parameters: 200, 50, 0.15) to identify and exclude individuals demonstrating extreme heterozygosity, specifically those with heterozygosity F coefficients exceeding three standard deviations from the cohort mean. To uphold data integrity, stringent filtering criteria were applied, removing SNPs and samples with call rates below 0.99 and SNPs exhibiting minor allele frequency (MAF) below 0.01. Sex-discrepancy filtering was also conducted to exclude participants whose genetically inferred sex conflicted with self-reported sex. Principal component analysis (PCA) was employed to assess and control for population stratification, facilitating the exclusion of non-European ancestry individuals by referencing HapMap populations. Relatedness filtering procedures were rigorously applied to eliminate potential cryptic relatedness, specifically removing individuals with first- or second-degree kinship (*π > 0.125*), thereby ensuring sample independence. Additionally, a haplotype-based test was utilized to detect and remove SNPs demonstrating significant non-random missing genotype data (*p < 1 × 10*⁻⁴). Hardy–Weinberg equilibrium (HWE) testing was subsequently conducted to exclude SNPs deviating significantly from equilibrium (*p < 1 × 10*⁻⁶), further reducing potential genotyping inaccuracies. Post-QC, genotype imputation was conducted using the Michigan Imputation Server, employing the Haplotype Reference Consortium (HRC) r1.1 2016 reference panel (hg19), significantly enhancing genomic coverage and imputation precision.

### Polygenic risk score for various mental disorder

The UK Biobank genetic dataset comprises 487,409 samples, which were phased and imputed to approximately 96 million variants using the Haplotype Reference Consortium (HRC) and the UK10K + 1000 Genomes reference panels ^27^.Samples with discrepancies between genetic and self-reported sex, extreme heterozygosity, or high genotyping missingness were excluded. Analyses were restricted to unrelated participants of European ancestry. At the variant level, SNPs were filtered based on imputation quality (INFO score < 0.3), minor allele frequency (MAF < 1%), missingness (>1%), and deviation from Hardy–Weinberg equilibrium in controls (p < 1 × 10⁻⁶). All genetic analyses were conducted using PLINK 1.9 ^56^ [https://www.cog-genomics.org/plink/1.9].

PRScs ^57^ [https://github.com/getian107/PRScs] and PLINK 2.0 [https://www.cog-genomics.org/plink/2.0/] were used to calculate polygenic risk scores (PRS). PRS were generated for eleven diseases using the most recent GWAS summary statistics: Parkinson’s disease (PD) ^58^, Alzheimer’s disease (AD) ^40^, Amyotrophic lateral sclerosis (ALS) ^59^, Lewy body dementia (LBD) ^60^, Anxiety ^61^, Alcoholism ^62^, Bipolar disorder (BP) ^63^, Insomnia ^64^, Major depressive disorder (MDD) ^65^, Post-traumatic stress disorder (PTSD) ^66^, Schizophrenia (SCZ) ^39^. PRS-CS applies a Bayesian regression framework with continuous shrinkage (CS) priors to estimate SNP effect sizes, and posterior SNP weights were inferred from each GWAS summary statistic. The European subset of the 1000 Genomes Project was used as an external linkage disequilibrium (LD) reference panel. Individual-level PRS were computed by summing posterior SNP weights across all chromosomes using PLINK’s --score command.

### The correlation between Polygenic risk score and structural asymmetric pattern

Furthermore, to evaluate differences between disease-related genes and asymmetry patterns, we computed correlations between individual loadings on the age-associated asymmetry patterns (i.e., those significantly related to age) and multiple disease polygenic risk scores (PRSs), controlling for age, sex, and handedness; p-values were corrected for multiple comparisons using FDR.

### The correlation between lifestyle behavior and structural asymmetric pattern

We further leveraged real-world wearable data to examine associations between aging-related asymmetry patterns and lifestyle factors. Specifically, we computed Pearson correlation coefficients (ρ) between asymmetry pattern scores and lifestyle phenotypes, with multiple comparisons controlled using FDR correction.

### Genetic mechanism

We deconvolved cell-type fractions from microarray profiles obtained from the Allen Human Brain Atlas (AHBA; http://human.brain-map.org/)^67^ to study how the asymmetric structural pattern of aging is regulated by genes. The AHBA dataset comprised 3,702 spatially discrete cortical samples from six post-mortem donors (mean age = 42.50 ± 13.38 years; 5 male, 1 female). Expression matrices were processed with the abagen toolbox ^68^(https://github.com/netneurolab/abagen) and assigned to Desikan–Killiany 68 cortical regions, after which regional expression patterns were correlated with aging patterns. Gene sets associated with significant expression–CT relationships were intersected with loci implicated by a large bipolar disorder genome-wide association study ^69^, yielding an overlap set for downstream analyses. We further tested whether genes showing transcriptional dysregulation in post-mortem cortex exhibited preferential expression in regions characterized by CT abnormalities. To contextualize convergent biology, genes linked to the most robust reorganization patterns were submitted to Metascape (https://metascape.org/gp/index.html#/main/step1) for pathway enrichment across >40 knowledge bases ^49^; enrichment was deemed significant at a false discovery rate of 5% using a null-model–based procedure.

## DATA AVAILABILITY

The clinical data could be accessed according to reasonable requests for corresponding authors. Neuromap (https://netneurolab.github.io/neuromaps/usage.html), ENIGMA toolbox (https://enigma-toolbox.readthedocs.io/en/latest/pages.html). Application number of UK biobank dataset is 619657.

## CODE AVAILABILITY

Code will be available on https://github.com/Laoma29/Publication_codes.

## Data Availability

All data produced in the present study are available upon reasonable request to the authors

## ACKNOWLEDGMENTS

Xiaobo Liu is supported by the China Scholarship Council. Yong Han received Henan AIMS Professional Development Fund [JBKY250316], and Henan Province science and technology research and development plan joint fund (industry) major project[235101610004]

---

**S-figure.1.**
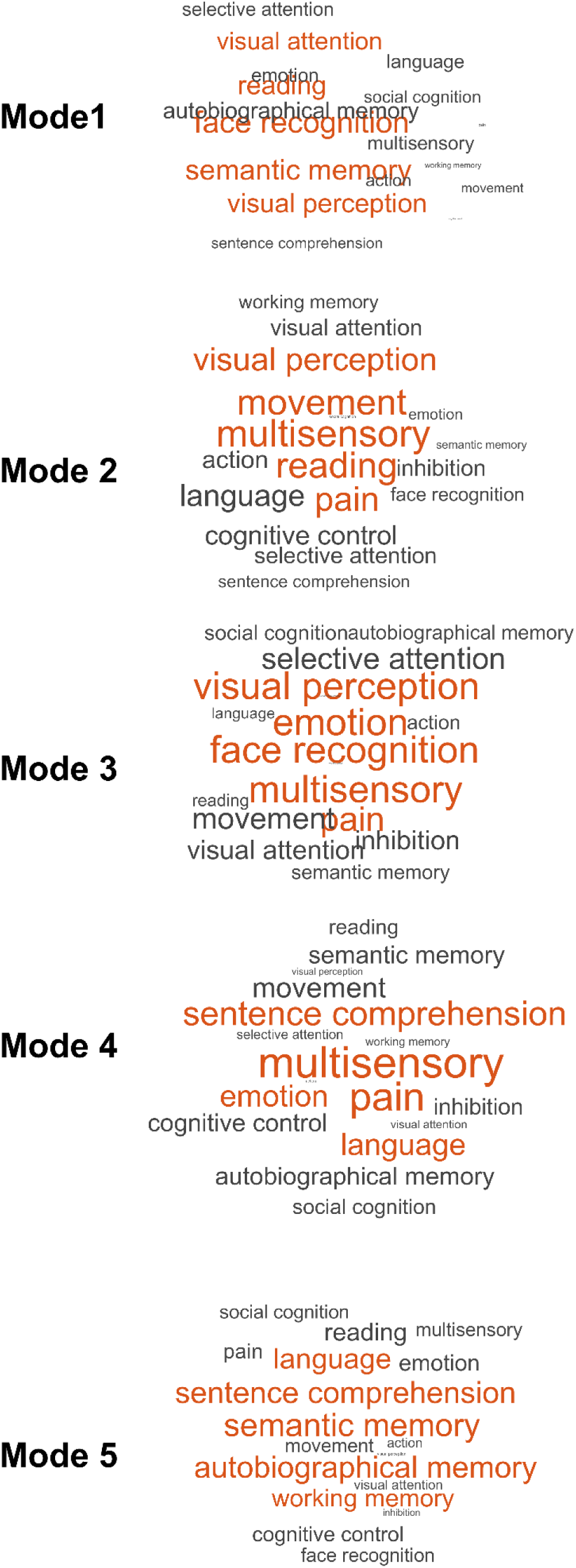
Cognition relevance of aging pattern.

## Reference

1. Roe, J. M. et al. Tracing the development and lifespan change of population-level structural asymmetry in the cerebral cortex. eLife 12, e84685 (2023).

2. Korbmacher, M. et al. Brain asymmetries from mid- to late life and hemispheric brain age. Nat. Commun. 15, 956 (2024).

3. Saltoun, K., Yeo, B. T. T., Paul, L., Diedrichsen, J. & Bzdok, D. Longitudinal changes in brain asymmetry track lifestyle and disease. Nat. Commun. 16, 5611 (2025).

4. Filippi, M. et al. Age-related vulnerability of the human brain connectome. Mol. Psychiatry 28, 5350–5358 (2023).

5. Zhang, R. et al. Brain age gap as a predictive biomarker that links aging, lifestyle, and neuropsychiatric health. Commun. Med. 5, 441 (2025).

6. Li, J., Lam, L. C. W. & Lu, H. Decoding MRI-informed brain age using mutual information. Insights Imaging 15, 216 (2024).

7. A review on brain age prediction models. Brain Res. 1823, 148668 (2024).

8. Madan, C. R. & Kensinger, E. A. Cortical complexity as a measure of age-related brain atrophy. NeuroImage 134, 617–629 (2016).

9. Yu, J. Age-related decline in thickness and surface area in the cortical surface and hippocampus: lifespan trajectories and decade-by-decade analyses. GeroScience 46, 6213–6227 (2024).

10. Li, M. et al. Changes in white matter functional networks across late adulthood. Front. Aging Neurosci. 15, (2023).

11. Dular, L., Špiclin, Ž., for the Alzheimer’s Disease Neuroimaging Initiative & the Australian Imaging Biomarkers and Lifestyle Flagship Study of Ageing. Analysis of Brain Age Gap across Subject Cohorts and Prediction Model Architectures. Biomedicines 12, 2139 (2024).

12. Petersen, M. et al. Brain network architecture constrains age-related cortical thinning. NeuroImage 264, 119721 (2022).

13. Lee, H. et al. Age-related alterations in human cortical microstructure across the lifespan: Insights from high-gradient diffusion MRI. Aging Cell 23, e14267 (2024).

14. Fujita, S. et al. Characterization of Brain Volume Changes in Aging Individuals With Normal Cognition Using Serial Magnetic Resonance Imaging. *JAMA Netw*. Open 6, e2318153 (2023).

15. Differences in the Lateralization of Theta and Alpha Power During n-Back Task Performance Between Older and Young Adults in the Context of the Hemispheric Asymmetry Reduction in Older Adults (HAROLD) Model. https://www.mdpi.com/2073-8994/16/12/1623.

16. Azzam, M. et al. A review of artificial intelligence-based brain age estimation and its applications for related diseases. Brief. Funct. Genomics 24, elae042 (2025).

17. van Es, M. W. J. et al. Large-scale cortical functional networks are organized in structured cycles. Nat. Neurosci. 28, 2118–2128 (2025).

18. Toga, A. W. & Thompson, P. M. Mapping brain asymmetry. Nat. Rev. Neurosci. 4, 37–48 (2003).

19. Lee, D. D. & Seung, H. S. Learning the parts of objects by non-negative matrix factorization. Nature 401, 788–791 (1999).

20. Zając-Lamparska, L., Zabielska-Mendyk, E., Zapała, D. & Augustynowicz, P. Differences in the Lateralization of Theta and Alpha Power During n-Back Task Performance Between Older and Young Adults in the Context of the Hemispheric Asymmetry Reduction in Older Adults (HAROLD) Model. Symmetry 16, 1623 (2024).

21. Kaufmann, T. et al. Common brain disorders are associated with heritable patterns of apparent aging of the brain. Nat. Neurosci. 22, 1617–1623 (2019).

22. Marquand, A. F., Rezek, I., Buitelaar, J. & Beckmann, C. F. Understanding Heterogeneity in Clinical Cohorts Using Normative Models: Beyond Case-Control Studies. Biol. Psychiatry 80, 552–561 (2016).

23. Reuter-Lorenz, P. A. & Park, D. C. How Does it STAC Up? Revisiting the Scaffolding Theory of Aging and Cognition. Neuropsychol. Rev. 24, 355–370 (2014).

24. Cole, J. H. & Franke, K. Predicting Age Using Neuroimaging: Innovative Brain Ageing Biomarkers. Trends Neurosci. 40, 681–690 (2017).

25. Bethlehem, R. A. I., et al. Brain charts for the human lifespan. Nature 604, 525–533 (2022).

26. Miller, K. L. et al. Multimodal population brain imaging in the UK Biobank prospective epidemiological study. Nat. Neurosci. 19, 1523–1536 (2016).

27. Bycroft, C. et al. The UK Biobank resource with deep phenotyping and genomic data. Nature 562, 203–209 (2018).

28. Ruigrok, A. N. V. et al. A meta-analysis of sex differences in human brain structure. Neurosci. Biobehav. Rev. 39, 34–50 (2014).

29. Ingalhalikar, M. et al. Sex differences in the structural connectome of the human brain. Proc. Natl. Acad. Sci. 111, 823–828 (2014).

30. Erickson, K. I. et al. Exercise training increases size of hippocampus and improves memory. Proc. Natl. Acad. Sci. 108, 3017–3022 (2011).

31. Xie, L. et al. Sleep Drives Metabolite Clearance from the Adult Brain. Science 342, 373–377 (2013).

32. Moderate alcohol consumption as risk factor for adverse brain outcomes and cognitive decline: longitudinal cohort study | The BMJ. https://www.bmj.com/content/357/bmj.j2353.

33. Morris, M. C. et al. MIND diet associated with reduced incidence of Alzheimer’s disease. Alzheimers Dement. 11, 1007–1014 (2015).

34. Orben, A. & Przybylski, A. K. The association between adolescent well-being and digital technology use. *Nat*. Hum. Behav. 3, 173–182 (2019).

35. Birney, E. Mendelian Randomization. Cold Spring Harb. Perspect. Med. 12, a041302 (2022).

36. Choi, S. W., Mak, T. S.-H. & O’Reilly, P. F. Tutorial: a guide to performing polygenic risk score analyses. Nat. Protoc. 15, 2759–2772 (2020).

37. Martin, A. R. et al. Clinical use of current polygenic risk scores may exacerbate health disparities. Nat. Genet. 51, 584–591 (2019).

38. Identification of risk loci with shared effects on five major psychiatric disorders: a genome-wide analysis. The Lancet 381, 1371–1379 (2013).

39. Trubetskoy, V. et al. Mapping genomic loci implicates genes and synaptic biology in schizophrenia. Nature 604, 502–508 (2022).

40. Bellenguez, C. et al. New insights into the genetic etiology of Alzheimer’s disease and related dementias. Nat. Genet. 54, 412–436 (2022).

41. Jansen, P. R. et al. Genome-wide analysis of insomnia in 1,331,010 individuals identifies new risk loci and functional pathways. Nat. Genet. 51, 394–403 (2019).

42. Rutherford, S. et al. The normative modeling framework for computational psychiatry. Nat. Protoc. 17, 1711–1734 (2022).

43. Martin, A. R. et al. Clinical use of current polygenic risk scores may exacerbate health disparities. Nat. Genet. 51, 584–591 (2019).

44. López-Otín, C., Blasco, M. A., Partridge, L., Serrano, M. & Kroemer, G. The Hallmarks of Aging. Cell 153, 1194–1217 (2013).

45. Sekar, A. et al. Schizophrenia risk from complex variation of complement component 4. Nature 530, 177–183 (2016).

46. Hawrylycz, M. J. et al. An anatomically comprehensive atlas of the adult human brain transcriptome. Nature 489, 391–399 (2012).

47. Richiardi, J. et al. Correlated gene expression supports synchronous activity in brain networks. Science 348, 1241–1244 (2015).

48. Arnatkevicčiūtė, A., Fulcher, B. D. & Fornito, A. A practical guide to linking brain-wide gene expression and neuroimaging data. NeuroImage 189, 353–367 (2019).

49. Zhou, Y. et al. Metascape provides a biologist-oriented resource for the analysis of systems-level datasets. Nat. Commun. 10, 1523 (2019).

50. Fischl, B. FreeSurfer. NeuroImage 62, 774–781 (2012).

51. Taylor, J. R. et al. The Cambridge Centre for Ageing and Neuroscience (Cam-CAN) data repository: Structural and functional MRI, MEG, and cognitive data from a cross-sectional adult lifespan sample. NeuroImage 144, 262–269 (2017).

52. Jernigan, T. L. et al. The Pediatric Imaging, Neurocognition, and Genetics (PING) Data Repository. NeuroImage 124, 1149–1154 (2016).

53. Altshuler, D., Donnelly, P., & The International HapMap Consortium. A haplotype map of the human genome. Nature 437, 1299–1320 (2005).

54. McCarthy, S. et al. A reference panel of 64,976 haplotypes for genotype imputation. Nat. Genet. 48, 1279–1283 (2016).

55. Das, S. et al. Next-generation genotype imputation service and methods. Nat. Genet. 48, 1284–1287 (2016).

56. Chang, C. C. et al. Second-generation PLINK: rising to the challenge of larger and richer datasets. GigaScience 4, s13742-015-0047–8 (2015).

57. Ge, T., Chen, C.-Y., Ni, Y., Feng, Y.-C. A. & Smoller, J. W. Polygenic prediction via Bayesian regression and continuous shrinkage priors. Nat. Commun. 10, 1776 (2019).

58. Program (GP2), T. G. P. G. & Leonard, H. L. Novel Parkinson’s Disease Genetic Risk Factors Within and Across European Populations. 2025.03.14.24319455 Preprint at 10.1101/2025.03.14.24319455 (2025).

59. van Rheenen, W. et al. Common and rare variant association analyses in amyotrophic lateral sclerosis identify 15 risk loci with distinct genetic architectures and neuron-specific biology. Nat. Genet. 53, 1636–1648 (2021).

60. Chia, R. et al. Genome sequencing analysis identifies new loci associated with Lewy body dementia and provides insights into its genetic architecture. Nat. Genet. 53, 294–303 (2021).

61. Forstner, A. J. et al. Genome-wide association study of panic disorder reveals genetic overlap with neuroticism and depression. Mol. Psychiatry 26, 4179–4190 (2021).

62. Zhou, H. et al. Multi-ancestry study of the genetics of problematic alcohol use in over 1 million individuals. Nat. Med. 29, 3184–3192 (2023).

63. Koromina, M. et al. Fine-mapping genomic loci refines bipolar disorder risk genes. Nat. Neurosci. 28, 1393–1403 (2025).

64. Watanabe, K. et al. Genome-wide meta-analysis of insomnia prioritizes genes associated with metabolic and psychiatric pathways. Nat. Genet. 54, 1125–1132 (2022).

65. Adams, M. J. et al. Trans-ancestry genome-wide study of depression identifies 697 associations implicating cell types and pharmacotherapies. Cell 188, 640–652.e9 (2025).

66. Nievergelt, C. M. et al. Genome-wide association analyses identify 95 risk loci and provide insights into the neurobiology of post-traumatic stress disorder. Nat. Genet. 56, 792–808 (2024).

67. Shen, E. H., Overly, C. C. & Jones, A. R. The Allen Human Brain Atlas: Comprehensive gene expression mapping of the human brain. Trends Neurosci. 35, 711–714 (2012).

68. Markello, R. D. & Misic, B. Comparing spatial null models for brain maps. NeuroImage 236, 118052 (2021).

69. Mullins, N. et al. Genome-wide association study of more than 40,000 bipolar disorder cases provides new insights into the underlying biology. Nat. Genet. 53, 817–829 (2021).

